# Alternative Substrates in the Critically Ill Subject (ASICS): Safety, Feasibility, Tolerability and Metabolic Profiling of a Novel Ketogenic Feed

**DOI:** 10.1101/2023.03.30.23287849

**Authors:** Angela McNelly, Anne Langan, Danielle E. Bear, Alexandria Page, Tim Martin, Fatima Seidu, Filipa Santos, Kieron Rooney, Kaifeng Liang, Tomas Baldwin, Simon J Heales, Isabelle Alldritt, Hannah Crossland, Philip J. Atherton, Daniel Wilkinson, Hugh Montgomery, John Prowle, Rupert Pearse, Simon Eaton, Zudin A. Puthucheary

## Abstract

Bioenergetic failure caused by impaired utilisation of glucose and fatty acids contributes to organ dysfunction across multiple tissues in critical illness. Ketone bodies may form an alternative substrate source, but the feasibility and safety of inducing a ketogenic state in physiologically unstable patients is not known.

Twenty-nine mechanically ventilated adults with multi-organ failure were randomised into a two-centre safety and feasibility trial of ketogenic versus standard enteral feeding. Ketogenic feeding was feasible, safe, well tolerated and resulted in ketosis. Patients receiving ketogenic feeding had fewer hypoglycaemic events (0% vs. 1.58%), required less exogenous insulin (0.0 IU (IQR 0-16) vs.78 IU (IQR 0-412) but had slightly more daily episodes of diarrhoea (53.5% vs. 42.9%) over the trial period. Untargeted metabophenotyping revealed altered Cahill cycle flux and bioenergetic states, suggesting an advantageous metabolic profile. Ketogenic feeding is feasible and may be a novel intervention for addressing bioenergetic failure in critically ill patients.

Clinical Trials.gov registration: NCT04101071; 19.09.2019.

**Take-home Message:** Critical illness leads to altered metabolic states and bioenergetic failure caused by impaired utilisation of glucose, fatty acids and amino acids. This contributes to organ dysfunction across multiple tissues. Ketones may provide a safe and acceptable alternative metabolic fuel enabling energy production and maintaining tissue homeostasis.

**Tweet:** Ketogenic enteral feeding in early critical illness is feasible, safe and may decrease insulin requirements.

## Introduction

Critical illness is a state of ill health with vital organ dysfunction and a high risk of imminent death if care (pharmacological or mechanical) is not provided and has the potential for reversibility^1^. The physiological characteristics of critical illness have significant overlap across a wide range of presenting diseases, challenging commonly used disease-related taxonomies^2^. Multiple diverse stressors result in a unifying state of altered tissue metabolism and bioenergetics, compounding organ dysfunction and cell death in multiple tissues such as the brain, lung, kidney and skeletal muscle^3-6^. Specifically, substrate utilisation in the tri-carboxylic acid (TCA) cycle is impaired in critical illness, with tissue hypoxia and inflammation prevent glucose-derived pyruvate from being converted to acetyl-CoA, as a result of the Pasteur effect^7 8^. Amino acids may be recycled for pyruvate reconstitution in starvation, but such processes (e.g., the Cahill cycle) are affected by tissue hypoxia, inflammation, impaired Glucose Transporter Type 4 (GLUT-4) translocation, exogenous insulin therapy and other hallmarks of critical illness^8-10^. Finally, mitochondrial fatty acid oxidation is downregulated, and the resultant inability to use any of these three substrates efficiently leads to a bioenergetic crisis^8 11 12^.

Under conditions of physiological stress, ketone bodies provide a source of substrate for ATP generation. In high intensity exercise, ketogenic diets provide ketone bodies for substrates, improving ATP production decreasing muscle protein breakdown and improving physical performance.^13^ During periods of starvation, brain metabolism relies on ketone bodies instead of fat or glucose^14^, and ketone bodies may provide up to 50% of total body basal energy, enabling the high-energy requirement of the human brain to be met whilst sparing muscle. ^15 16^ Ketogenic diets reverse the metabolic defects of non-alcoholic fatty liver disease.^17^ Patients with diabetes use ketone bodies for cardiac ATP synthesis.^18^ Ketone bodies such as beta-hydroxybutyrate and acetoacetate are the result of hepatic metabolism of fatty acids. Ketolysis occurs in mitochondria of extra-hepatic tissues, resulting in the formation of acetyl-CoA. The rate-determining step is the reconstitution of acetoacetyl-CoA from acetoacetate by the enzyme succinyl CoA-oxoacid transferase, which is not regulated by hypoxia or inflammation, unlike pyruvate dehydrogenase kinase^19^. Ketone bodies may therefore offer an alternative substrate source for energy production in critically ill patients.In addition, ketones may have other beneficial impacts in critically ill patients: in those with Acute Respiratory Distress Syndrome (ARDS), beta-hydroxybutyrate metabolically reprogrammes T-cells to improve functionality.^20^

However, the feasibility and safety of achieving ketosis in unstable patients in multi-organ failure has yet to be proven. Ketoacidosis might occur if ketones were not metabolised, exacerbating pre-existing systemic and cellular acidosis that carries a mortality risk to critically ill patients. Ketogenic diets minimise exogenous glucose delivery, which might predispose patients to hypoglycaemia, which is harmful to patients^21^. Lastly, a ketogenic high lipid feed might increase the risk of vomiting (and therefore pulmonary aspiration), diarrhoea and pancreatitis ^22^.

One specific physiological consequence of critical illness that may be attenuated by a ketogenic diet is that of muscle wasting. Critically ill patients lose 2-3% of their muscle mass per day^23^. This is associated with increases in length of stay and mortality, and associated physical functional disability may persist for up to 5 years^24^. Patients, carers and health services are burdened by this physical function disability, which is recognised as a public health issue^25 26 27^. Muscle wasting and subsequent weakness in these patients has proven resistant to all forms of exercise rehabilitation and increased nutritional delivery of energy and protein^28 29^. This loss of muscle mass appears driven by a decrease in muscle protein synthesis and unchecked muscle protein breakdown ^23^. This in turn may be a consequence of bioenergetic failure and a lack of adenosine tri-phosphate production^8^. Muscle protein synthesis is a highly energy-dependent process and is likely to remain depressed until muscle bioenergetics normalise.

We therefore performed a randomised trial to determine the feasibility and safety of delivering a ketogenic enteral feed in critically ill patients and collecting physical function specific outcomes. We additionally performed an exploratory analysis of plasma metabolomic profiling to ascertain the presence or absence of a signal for efficacy in altering tissue metabolism, which might warrant further research in a larger trial.

## Methods

We performed a single-blinded randomised controlled feasibility trial in two UK intensive care units (ICUs), with an allocation ratio of 1:1.

### Participants

Adult (≥18 years) ICU patients were screened for inclusion on weekdays, being eligible for enrolment up to 48 hours after ICU admission.

#### Inclusion Criteria

Requiring enteral nutrition via nasogastric tube; expected to be intubated and ventilated ≥48 hours; multi-organ failure (Sequential Organ Failure Assessment (SOFA) score >2 in >2 domains)^30^; likely ICU stay ≥5 days and likely survival ≥10 days (assessed as previously by senior ICU clinicians^23^).

#### Exclusion Criteria

Primary neuromyopathy or significant neurological impairment at the time of ICU admission that would preclude physical activity; unilateral/bilateral lower limb amputation; requirement for sole or supplementary parenteral nutrition; need for specialist nutritional intervention; known inborn error of metabolism; participation in another clinical trial. Patients at risk of refeeding syndrome (based on NICE guidelines^31^) were assessed on an individual basis.

Prospective informed assent was by nominated personal consultee (in person or by telephone) or professional consultee. Retrospective participant consent was obtained on return of each participant’s mental capacity. Permission to use participants’ data if capacity did not return or if they did not survive, was included in the assent process.

The study received ethics committee approval (National Research Ethics Service Committee Wales 5 – Bangor; REC reference 19/WA/0209; IRAS project ID 266031) and was publicly registered prior to the first patient being randomised (ClinicalTrials.gov, NCT04101071). We used the CONSORT (Consolidated Standards of Reporting Trials) statement when reporting this trial^32^.

### Feeding Regimens

The ketogenic enteral feed was reconstituted for each patient in a clean kitchen area of the ICU by research nurses, with the proportions of individual nutritional components used devised by a dietitian using K.Quik® (Vitaflo International Ltd, Liverpool, UK), Renapro® (Stanningley Pharma, Nottingham, UK), Maxijul® (Nutricia, Liverpool, UK) and Fresubin® 5kcal (Fresenius Kabi, Dublin, Ireland, if additional fat was needed). Ketogenic and standard enteral feeding regimens were provided continuously as per the standard protocol for each trust.

Patients were ineligible if they received ≥12 hours of standard feed prior to randomisation. A dietitian assessed individual patients’ nutritional needs within 72 hours of randomisation. The Modified Penn State equation or a weight-based equation (e.g., 25 kcal/kg) was used to estimate energy targets. Protein targets were individualised to each patient with a range of 0.83 -1.5g/kg/d being used according to specific clinical need. Patients were considered to have received adequate nutrition if they achieved >80% of their prescribed targets. Ketogenic enteral feeding continued for the duration of the 10-day trial period as tolerated, before reverting to standard enteral feed, as per the clinician responsible for the patient’s care. Patients in the control arm received the site-specific enteral feed as per Trust protocols with an agreed daily energy target to meet their nutritional needs. Multivitamins were administered daily in the ketogenic arm as micronutrients were otherwise not in the modular feed. Intravenous glucose was only to be administered for the emergency treatment of hypoglycaemia. Blood glucose levels in both feeding arms were managed according to local protocols.

### Endpoints

The primary endpoints were patient recruitment and consent rates during screening (with retention rates); the ease of reconstitution and administration of the novel ketogenic enteral feed by ICU and research staff (determined via questionnaire; safety (reports of adverse events [AEs] and serious adverse events [SAEs]); parameters of enteral feed absorption and blood chemistry (including glucose levels and achievement of ketosis) post-recruitment; and plasma concentrations of beta hydroxybutyrate and acetoacetate, glucose, lactate, pyruvate and medium-chain fatty acids from blood samples at timepoints during the 10-day study period (See Table 1)

**Table 1:** Feasibility and safety outcome measures. ICU=Intensive Care Unit, NOK=Next of kin, RF_CSA_=Rectus femoris muscle cross sectional area, CPAx=Chelsea Critical Care Physical Assessment Tool, 6-MWD=6-Minute Walk Distance, SPPB=Short Physical Performance, GCMS=Gas Chromatography Mass Spectrometry, HPLC=High-Performance Liquid Chromatography.

| ENDPOINT | TOOL/METHOD (REFERENCE) | OBJECTIVE |
| --- | --- | --- |
| **A) Patient recruitment** | From screening clinical records, medical history, demographic information; informed consent; randomisation | Feasibility of recruiting to a trial of ketogenic enteral feeding |
| **B) Modular feed preparation (on ICU) to meet dietician-prescribed nutritional targets** | Staff questionnaire | Feasibility of preparation of modular feed on ICUs |
| **C) Administration of feed** | Staff questionnaire | Feasibility of giving ketogenic feed |
| **D) Glucose and lactate measurements** | Point-of-care tests; routine biochemistry | Feasibility of glucose and lactose control |
| **E) EQ-5D-5L questionnaire** | Completed by NOK proxy either in-person or via telephone | Feasibility of determining quality of life at baseline, 3-, 6- and 12 months |
| **F) Energy and protein intake** | From feed recipe and volume administered | Delivery of adequate energy and protein target |
| **G) Blood gases; biochemistry** | Routine biochemistry analysis of blood and urine samples | Safety of ketogenic feed |
| **H) Serious Adverse Events/Adverse Events** | Measures of Gastric intolerance: Gastric Residual Volume; diarrhoea; vomiting; use of pro-kinetics; daily blood glucose levels | Feeding intolerance; abnormal glucose control; other Adverse Events |
| **I) Muscle mass** | RF<sub>CSA</sub> measurements | Feasibility of bedside ultrasound scans |
| **J) Functional tests** | Physiotherapy assessments: CPAx; 6-MWD; SPPB | Effect on physical function |
| **K) Levels of ketone bodies,; pyruvate,; medium/long chain fatty acids; metabolite abundance** | GCMS and HPLC analysis of plasma samples | Induction of ketosis within study period; effect on nutrient metabolism |
| **L) blood urea levels in plasma** | Plasma samples | Effect on protein metabolism |
| **M) Indirect calorimetry** | Calorimeter | Effect on basal metabolic rate |
| **N) Completion of CRF** | Review of electronic medical records | Feasibility of collecting data |

Secondary endpoints included arterial blood gas parameters, ultrasound-determined rectus femoris cross-sectional area (as a marker of muscle loss); clinical, physiological and nutritional data; non-invasive metabolic data via indirect calorimetry; measures of physical function and quality of life; employment status and health care resource usage; and urinary concentrations of beta-hydroxybutyrate and total nitrogen, and plasma metabolomics at timepoints during the 10-day study period.

Measures of adverse safety impacts included daily rates of vomiting (>10mls); number of episodes of diarrhoea (Bristol Stool Score ≥5^33^ on 3 or more days) and daily rates of diarrhoea; daily rates of high gastric residual volume (GRV≥300ml), or impaired glycaemic control. Normoglycaemia was defined as a blood glucose concentration of 4-10mmol/l, and thus concentrations of ≥10.1 or ≤3.9mmol/l as hyperglycaemia or hypoglycaemia respectively. Details of longer-term secondary endpoints collected are provided in the Supplementary Information.

### Blood and Urine analyses

Plasma medium chain fatty acids, lactate, and beta-hydroxybutyrate, whole blood acetoacetate pyruvate, and urinary beta-hydroxybutyrate were analysed by gas chromatography/mass spectrometry^34^.

### Ultra High-Performance Liquid Chromatography Untargeted Metabolomics

Metabolites were extracted from plasma using a dual phase Bligh-Dyer extraction and analysed using a combination of hydrophilic interaction liquid chromatography (HILIC) and Reverse Phase Ultra High-Performance Liquid Chromatography-mass spectrometry (UHPLC-MS) for polar and non-polar/lipid metabolites respectively^35^. Metabolite features were extracted from raw files using XCMS, then quality-control filtered and normalised according to standard procedures. Validated metabolite features were annotated to provide putative metabolite ID’s. Metabolites were putatively identified by metID which utilises m/z and MS^2^ spectra matching from public metabolomics databases^36^. Mapping of metabolites to relevant pathways was performed by over representation analysis (ORA) using MetaboAnalyst^37^. Pathway Impact scores represent an objective estimate of the importance of a given pathway relative to the global metabolic network^38^. A cut off value of 0.1 was used, in keeping with previous work across multiple comparisons to filter less important pathways^38 39^.

Full details of sampling and analytical methods are provided in the Supplementary Information.

### Sample Size

Sample sizes of 12 per arm have been recommended where previous data on which to base a power calculation are lacking^40^. We aimed to recruit at least 37 patients to allow for a possible high drop-out rate from early death and early recovery, and for protocol violations (common in many critical care trials), and to thus leave12 patients per arm.

### Randomisation and Blinding

Randomisation was stratified for recruitment site (1:1 basis) and occurred once assent was obtained. Treatment arm allocation used an independent remote electronic web-based random allocation service to generate an unpredictable allocation outcome, and to conceal that outcome from research staff until assignment occurred. Whilst patients were blinded (by virtue of illness), knowledge of the feeding intervention they were receiving would not influence outcome. Where possible individuals collecting outcome assessments were blinded in this feasibility study, but not those preparing and administering feed due COVID-19-related staffing limitations. DB (who assessed all ultrasound scans) was masked to allocation until data analysis was complete (see Supplementary Information).

#### Statistical Analyses

Descriptive analysis was performed for the continuous outcomes using mean (95% confidence intervals) and Student’s T-test, or median (range) analysed using Mann Whitney U test. Chi-squared testing was used for proportional data. Recruitment rates are shown as a percentage with 95% confidence interval. All analyses were performed on an intention-to-treat basis using GraphPad (Prism) Two-tailed tests were used, and statistical significance was indicated by P≤0.05.

For metabolomic analyses, data were centred and scaled to perform sparse partial least squares discriminant analysis (SPLS-DA) validated by k-fold cross validation to establish differences between ketogenic and control diet groups at baseline and on day 10. Owing to the high variability in this data set, orthogonal projection to latent structures (OPLS) was utilised to maximise variation^41^. Models with error rate greater than 20% were considered to be overfitted (i.e., the model described random error in the data rather than relationships between variables)^42 43^. In non-overfitted models, all variables with a Variable Importance Projection (VIP) score greater than 1 were retained for further analysis^44^. The VIP score is a quantitative assessment of the discriminatory power of each individual feature^45^.

For time course analysis, a linear mixed effect model was fitted to each data matrix using the limma package^46^. Diet and time were fixed effects. Participant ID was a random effect to account for subject specific variation. Contrast matrices were set up comparing metabolite abundance at baseline and days 3, 5 and 10 of the intervention. Empirical Bayes moderated t-tests were performed to obtain p-values. False discovery rate (FDR) was accounted for using the Benjamini-Hochberg procedure. Metabolites were deemed significantly different between comparisons when FDR<0.05.

Further details on study methodology are available in the Supplementary Information.

#### Data Availability Statement

The data that support the findings of this study are available for research purposes on reasonable request from the corresponding author [ZAP]. The data are not publicly available since this was not included in research participant consent.

## Results

### Safety, Feasibility and Tolerability

Participants were recruited between 26th September 2019 and 22nd April 2021 (including two COVID-19-pandemic related pauses) from two United Kingdom ICUs. The CONSORT flow chart is available in the Supplementary Information (Figure S1). A total of 293 patients were screened, with 29 patients randomised after meeting inclusion criteria (see Supplementary Information Table S3) and a refusal rate for assent of 4.1% (12 patients). The rate of recruitment was 2.2 participants/month for the 13 months enrolment period (Figure S2). Participant retention rate was 82.8% (24 patients). Reasons for withdrawal are shown in Supplementary Information Table S4.

Participant demographics are shown in Table 2: mean (95%CI) APACHE II score was higher in the control arm than the ketogenic feeding arm (21.6 (18.4-24.8) vs. 16.4 (13.5-19.3); p=0.025), admission SOFA scores were similar (9.9 (95%CI8.4-11.4) vs. 10.1 (8.7-11.6); p=0.621). At the end of intervention period, the number of days of vasopressor support and total daily propofol dose were also higher in the control arm.

**Table 2:**
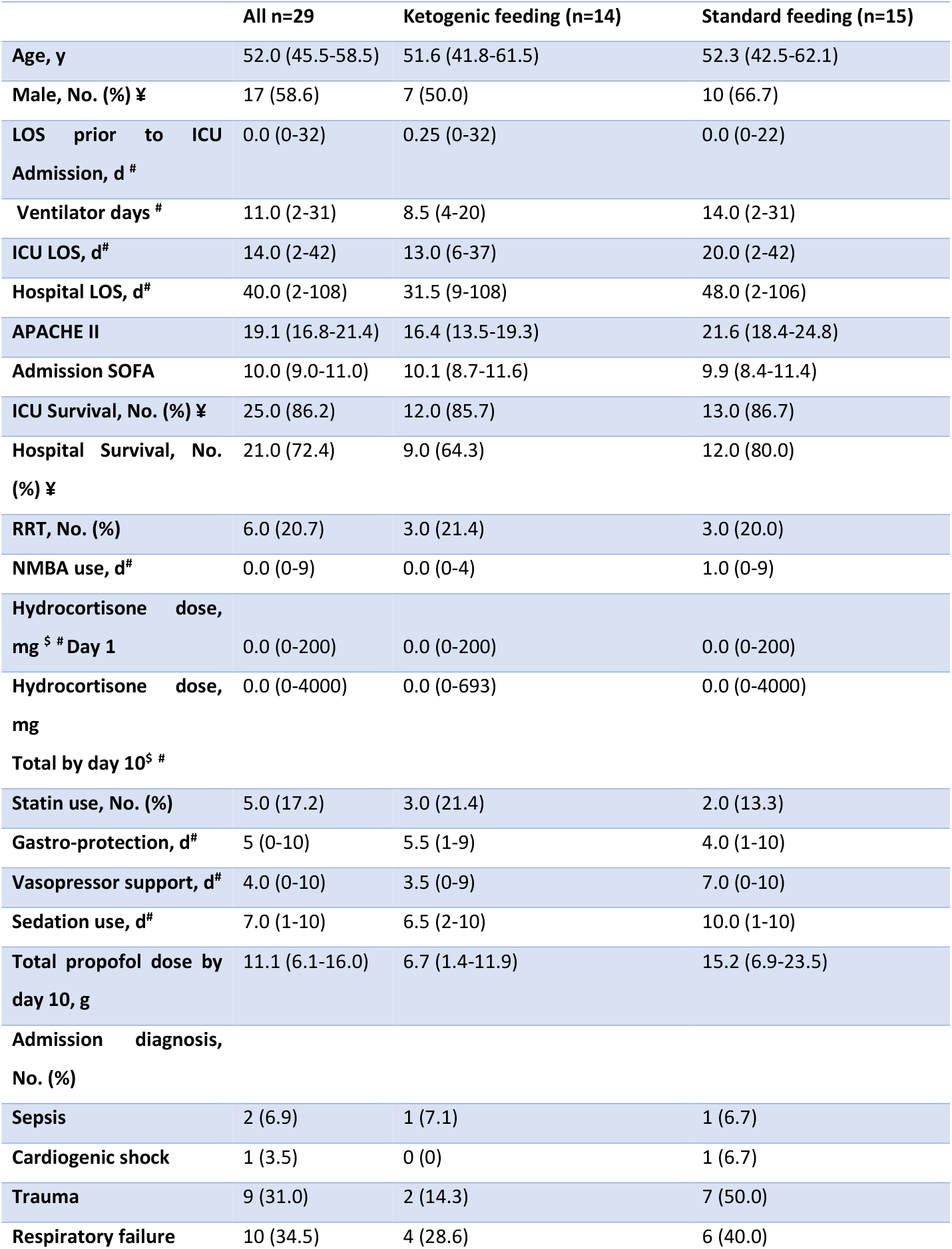
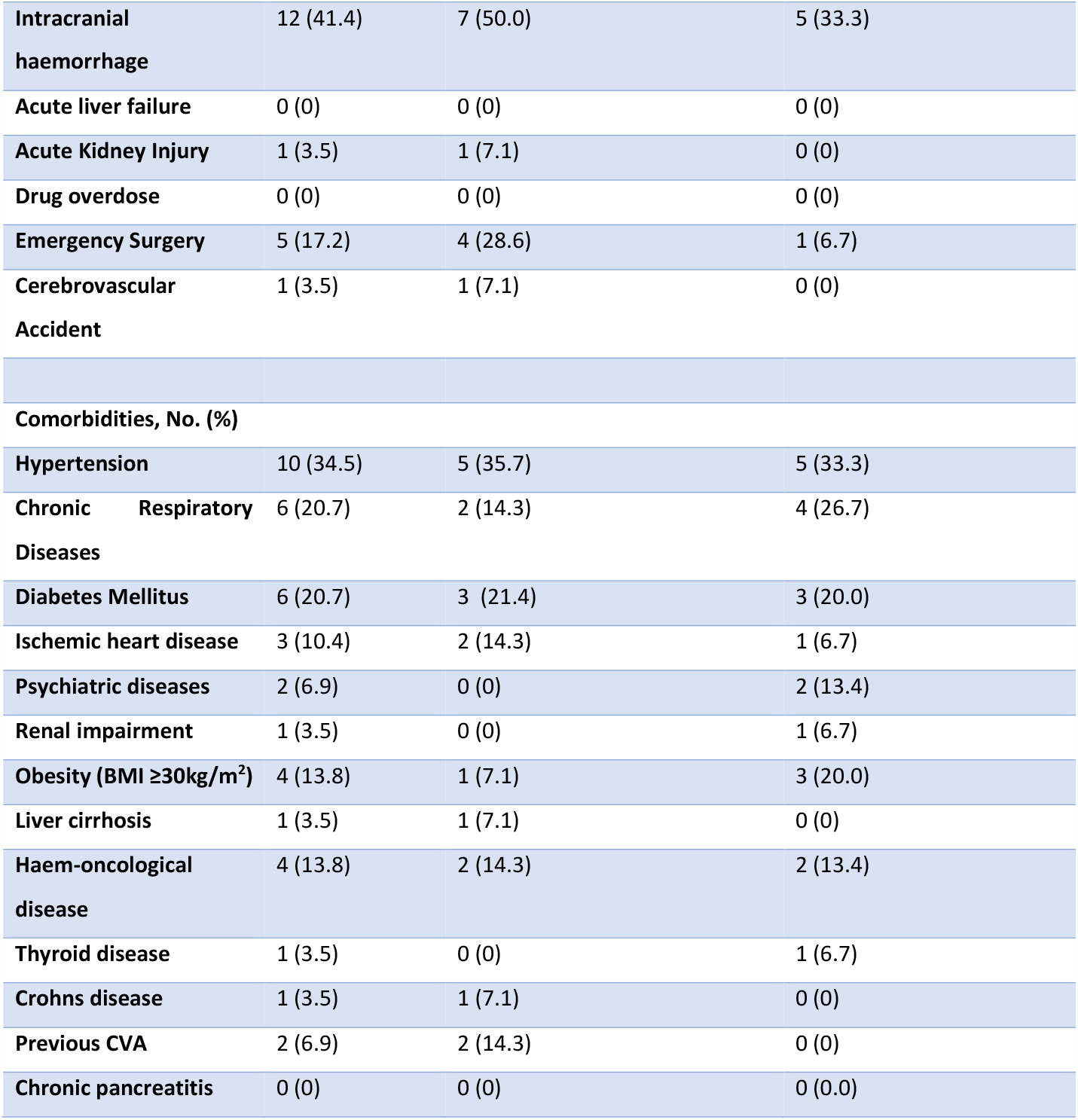
Patient characteristics and demographics. ICU=intensive care unit, APACHE II=Acute Physiology and Chronic Health Evaluation score, y-year, d=day, No.=number. LOS=Length of Stay, RRT=Renal Replacement Therapy, NMBA=Neuromuscular Blockade Agent; CVA=Cerebrovascular Accident $=Corticosteroid dosing as hydrocortisone equivalents. Data are mean (95% Confidence Intervals), except for # indicating median with range. Student’s T-test was used except for ¥ (Chi-squared) and # (Mann Whitney U).

#### Process and Feasibility of Nutritional Delivery

All patients randomised to the intervention received ketogenic enteral feeding. Feedback was obtained from 23 staff (4 research nurses, 16 ICU nurses, 1 pharmacist, 1 dietitian, 1 ICU consultant). The trial process was considered acceptable and feasible (See Supplementary Information Figure S3), although the preparation of the modular feed was considered laborious. A mean score of 8/10 (with 10 scored as the most positive response) was obtained for the question ‘*How keen would you be to work on another similar study*?’

#### Serious Adverse Events

Four serious adverse events (SAEs) were reported, and all were deemed to be unrelated to the intervention. Details of these are available in the Supplementary Information (Table S5). No episodes of pulmonary aspiration were reported.

#### Adverse Events

Similar proportions of gastrointestinal events were reported between arms, with the exception of diarrhoea. The proportion of patients with diarrhoea was greater in the ketogenic enteral feeding arm (intervention vs. control 76.9% vs. 52.3%) but the difference in proportion of daily episodes less marked (53.5% vs. 42.9%). One patient in the intervention arm was transferred to total parenteral nutrition on Day 6 as a result of concerns regarding enteral feeding intolerance (Supplementary Information Table S6).

Mean base excess and bicarbonate level were similar between arms, remaining within the normal ranges (+2 to -2) and (22mmol/L – 29mmol/L) respectively. One patient from each arm developed Acute Kidney Injury (Supplementary Information Table S7).

#### Development and Establishment of Ketosis

Ketosis was achieved within 48 hours and sustained for the 10-day intervention period, (Figure 1AB and Supplementary Information Figure S4). Medium chain fatty acid (octanoic acid and decanoic acid) concentrations from the ketogenic feed were higher in the intervention arm. As expected, no differences were seen between arms in dodecanoic acid concentrations, which were not part of either feed (Supplementary Information Figure S5). As a result, the ratio of octanoic acid to dodecanoic acid (C8+C10:C12) was higher in the intervention arm over time (Supplementary Information Figure S5D).

**Figure 1AB:**
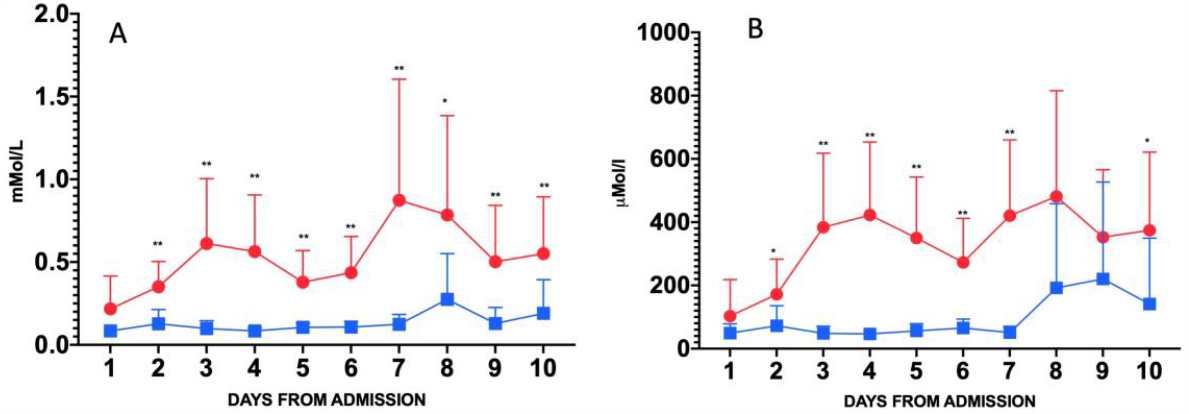
Plasma Beta-hydroxybutyrate (A) and Acetoacetate (B) concentrations during the 10-day intervention. Data are mean (95%CI) Red lines represent ketogenic feeding, and blue lines controls. *p<0.05; ** p<0.01 for two-tailed Mann-Whitney-U test.

#### Glucose Control

Two hypoglycaemic events were reported in the control arm and none in the intervention arm (1.58% vs 0% respectively). Hyperglycaemia occurred in fewer patients in the ketogenic enteral feeding arm (intervention vs. control 26.85% vs. 57.48%). In keeping with this, the coefficient of variation of daily glucose was lower in the intervention arm (9.4% vs. 14.8%, Figure 2A) as was median (IQR) cumulative insulin use (0.0 IU (IQR 0-16) vs.78 IU (IQR 0-412).

**Figure 2ABCD:**
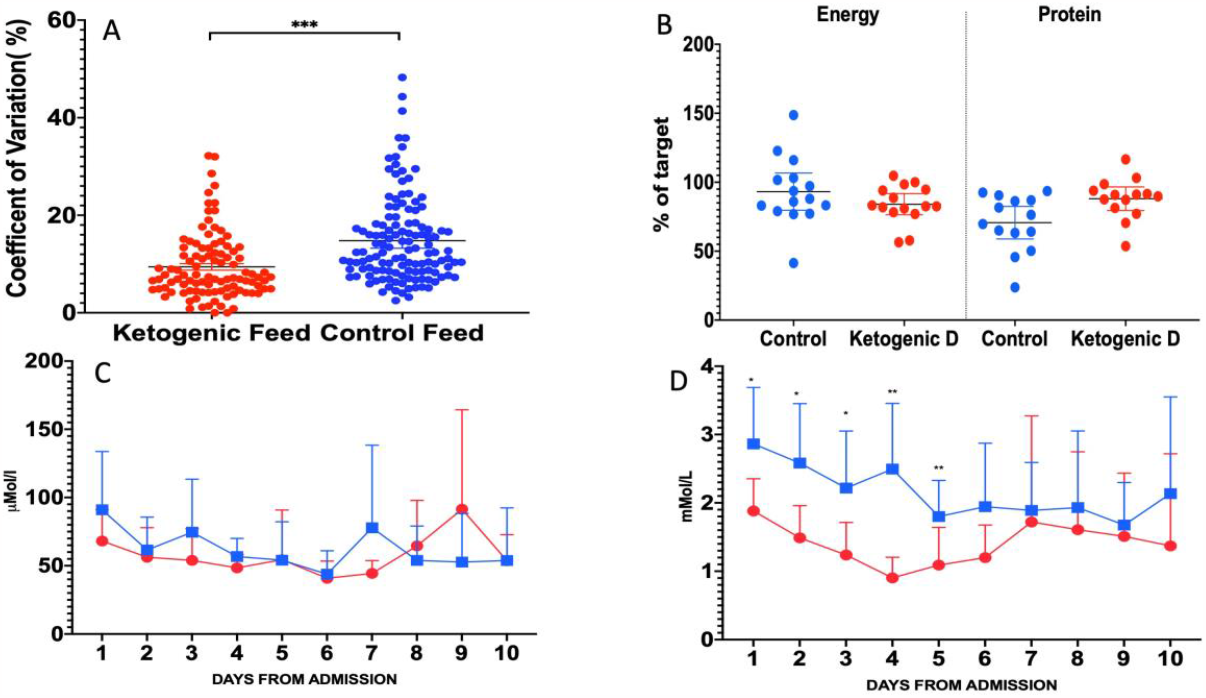
Co-efficient of Variation of serum glucose (A); Nutritional adequacy (B); Plasma pyruvate (C) and Lactate (D) concentrations during the intervention. Data are mean (95%CI). Red represents ketogenic feeding, blue controls. *p<0.05; ** p<0.01; *** p<0.001. between arms (Mann Whitney-U test).

#### Nutritional Adequacy and Substrate Utilisation

In per-protocol analysis, participants receiving control enteral feeding met 90.4% and 79.3% of energy and protein targets respectively; patients receiving ketogenic enteral feeding received 83.3% and 84.4% of energy and protein targets respectively (Figure 2B). This was similar in the Intention-To-Treat group, although a lower proportion of control arm participants met their protein target compared with the ketogenic enteral feeding arm (71.4% vs 88.1%). Indirect calorimetry was performed on a subset of patients. RQ was 0.83 in the control patients (n=6) and 0.78 in the intervention arm (n=8) (Supplementary Information Figure S6).

Plasma pyruvate concentrations were similar between arms (Figure 2C). Plasma lactate concentrations were lower in the ketogenic enteral feeding arm at baseline and remained lower throughout the study period Figure 2D). Collection of 24-hour urine samples to obtain total nitrogen values was not feasible in the context of heightened infection control during the pandemic.

#### Data Collection Completeness

The completion rate of data collection (for blood gases, biochemistry, haematology, bedside physiology, nutritional data and propofol usage) from medical records into the electronic database was 98.7% for those participants still in the study.

#### Rectus Femoris Ultrasound

Twenty-seven (93.1%) patients had 73 ultrasound scans performed over the period of the study. Scans were not performed in 2 (6.9%) of patients due to transfer to palliative care before the Day 1 scan was performed. However, scan quality did not reach acceptability in scans on 61 days (in 25 patients) when examined independently (DB and ZP). Reasons for this included the turnover and disruption of shift patterns of research nurses during the pandemic, and difficulties of working in protective equipment (PPE) leading to issues with consistent training and quality control.

#### Physical Functional Outcomes

The Chelsea Critical Care Physical Assessment Tool (CPAx) was completed in 26/29 (90%) patients at ICU discharge and 17/29 (59%) at hospital discharge. Collection of data for other physical function milestones (e.g. sit-to-stand 20.8%, bed-to-chair 29.2%, Short Physical Performance Battery 9.5%, 2 and 6 minute-walk tests <5%) was severely impeded by COVID-19-related limitations in access to physiotherapists for these assessments due to re-deployment.

#### Longer-term Outcomes

Quality of life assessed by EQ-5D-5L was measured at 3-, 6- and 12-months post-ICU discharge in 21 (72.4%), 21 (72.4%) and 19 (65.5%) of study participants respectively (including data from those who had died where available). Three non-responders needed a translator that was not available during the pandemic, two were in long-term care, and one had moved abroad. Primary healthcare usage data were available in 20/29 (69%) of patients. Questions of employment were abandoned due to the complexity of employment status during the COVID-19 pandemic, and the potential to cause patient distress. Final discharge location data were completed for 15/29 patients (51%).

### Metabolomic Analysis

Plasma metabolomic analyses identified a total of 185 metabolite features.

#### Exploratory Visualisations

Sparse Partial Least Squares Discriminant analysis (SPLS-DA) demonstrated no difference in metabolite abundance between arms on Day 1 (Supplementary Information Figure S7). By the end of the intervention period, between-arm differences were seen in 31 non-polar negative and 67 non-polar positive metabolites with Variable Importance in Projection (VIP) scores of >1. Similarly, 45 polar negative and 65 polar positive metabolites had a VIP score >1 (Figure 3ABCD).

**Figure 3ABCD:**
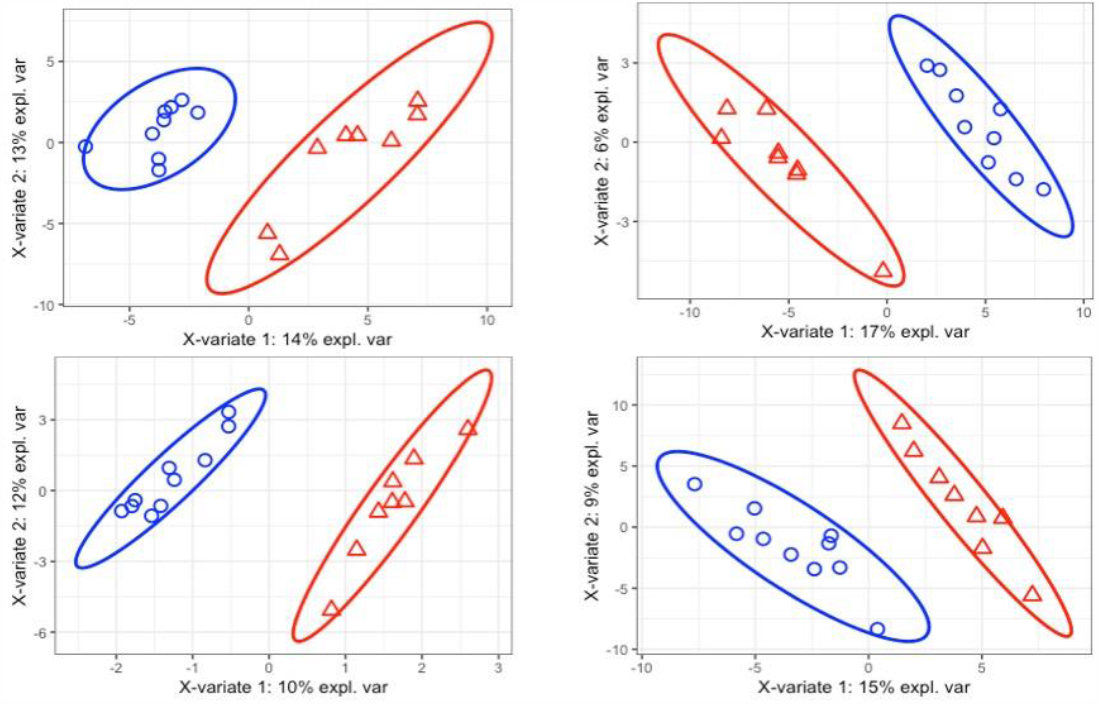
Sparse Partial Least Squares Discriminant analysis of patient randomised to ketogenic feeding on Day 10 (red triangle) and control feeding on day 10 (blue sphere). Clockwise from top right: polar positive, non-polar positive, non-polar negative, polar negative. Error rates are <20% (12%, 15%, 14% and 17% respectively), suggesting plot is a result of real variation.

Each arm additionally demonstrated changes (VIP scores>1) in metabolite abundance over time. In the control arm, 37 non-polar negative, 39 non-polar positive, 41 polar negative and 18 polar positive metabolite abundances were differentially altered over time (Figure S8). In the ketogenic enteral feeding arm, 23 non-polar negative, 22 non-polar positive, 59 polar negative and 20 polar positive metabolite abundances were differentially altered over time (Figure S9).

#### Pathway Analysis

Metaboanalyst pathway analysis demonstrated differential metabolite abundance in ketogenic enteral feeding arm vs. controls in beta-alanine metabolism (Impact 0.5), glycerophospholipid metabolism (Impact 0.2) and pentose and glucoronate interconversions (Impact 0.14).

Over time, changes in metabolite abundance in the ketogenic enteral feeding arm were seen in pantothenate and CoA biosynthesis (Impact 0.1) and alpha-linoleic acid metabolism (Impact 0.33). This was different to that seen in the control arm of caffeine metabolism (Impact 0.69), terpenoid backbone synthesis (Impact 0.11) and pentose and glucoronate interconversions (Impact 0.14). These data, and non-impactful pathways are summarised in Table 3.

**Table 3:**
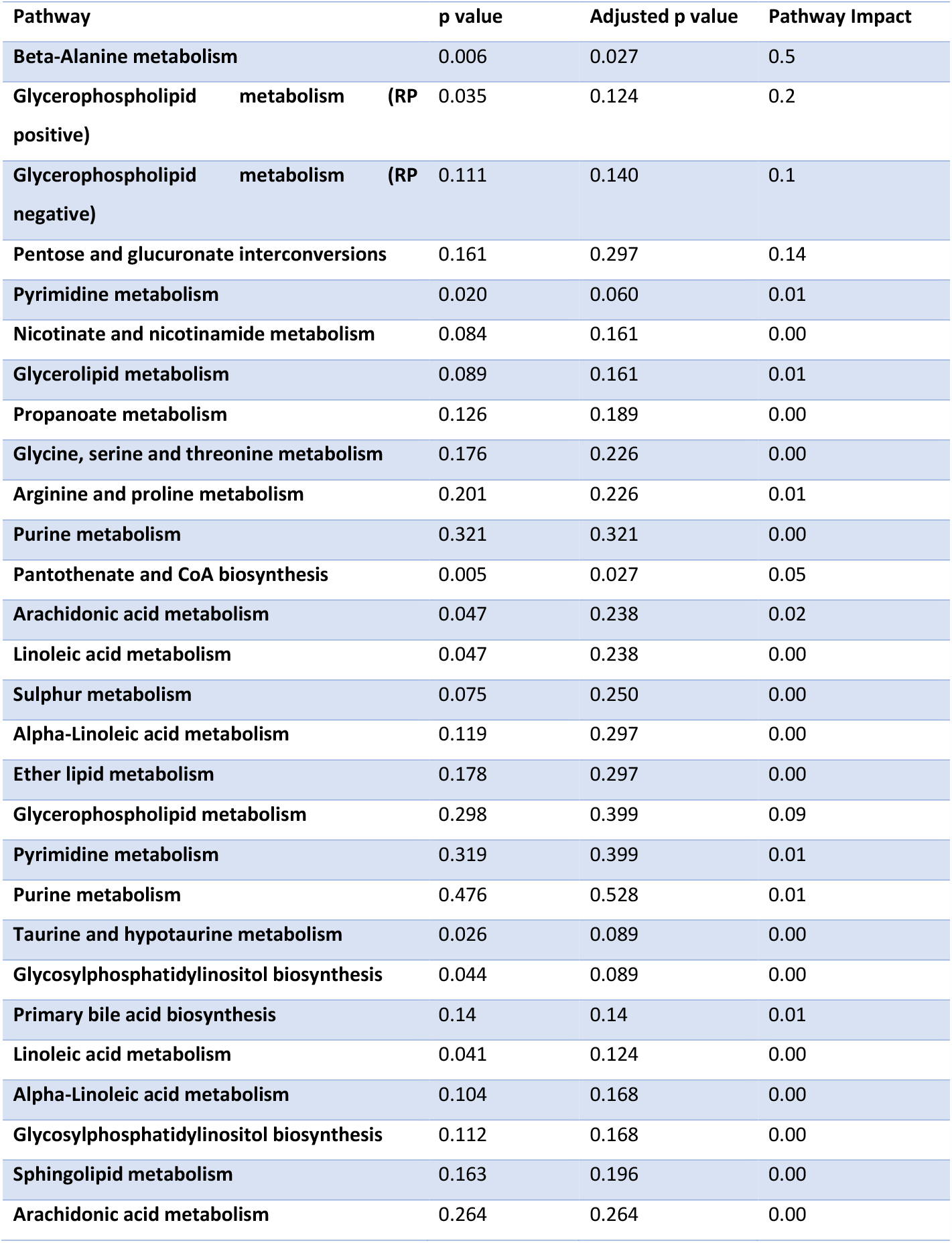
Pathway Impact Scores following mapping of metabolites to relevant pathways. Performed by over representation analysis (ORA) using MetaboAnalyst. Pathway Impact scores represent an objective estimate of the importance of a given pathway relative to the global metabolic network^38^. A cut off value of 0.1 was used, in keeping with previous work across multiple comparisons to filter less important pathways^38 39^.

#### Specific metabolite alterations

Specific metabolites driving the differential pathway abundance data were then examined. The ketogenic enteral feeding arm demonstrated increases in beta-alanine (17.2AU (17.0-17.4) vs 16.3AU (16.0-16.6); p=0.008) and ureidopropionic acid (16.1AU (15.7-16.4) vs 15.2AU (14.8-15.6); p=0.008) abundance and decreases in lithocholate 3-0-glucuronide (14.9 AU(14.8-15.0) vs 15.3AU (15.2-15.4); p=0.0004) relative to the control feed. Differences in glycerophospholipid metabolism was driven by changes in phosphocholine residues (Figure 4ABCDEFGH).

**Figure 4ABCDEFGH:**
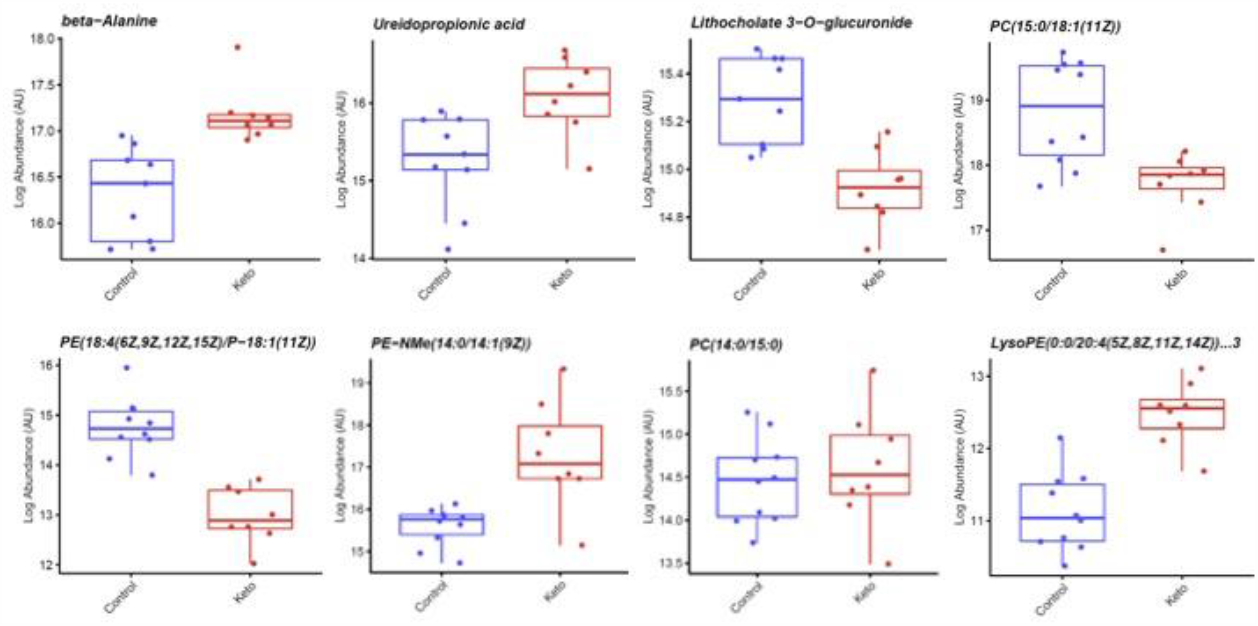
Log abundance in Arbitrary Units (AU) of metabolites driving differential pathway analysis. Data are mean (95%CI).

Over time, administration of ketogenic enteral feed was associated with a difference in ureidopropionic acid abundance (16.1AU (15.7-16.4) at day 10 vs. 15.0AU (14.7-15.2) at day 1; p=0.0002).In the control feed arm, differences over time in paraxanthine (15.0AU (14.3-15.6) at day 10 vs 15.9AU (15.3-16.5) at day 1; p=0.04), palmitoyl glucoronide (16.3AU (16.0-16.6) at day 10 vs 15.7AU(15.5-15.9) at day 1; p=0.003) and mevalonic acid (14.5AU (13.7-15.2) at day 10 vs 15.9AU (15.0-16.7) at day 1; p=0.02) were noted.

## Discussion

We have demonstrated that inducing sustained ketosis using a ketogenic enteral feeding regimen is safe, well tolerated, and feasible in critically ill patients with multi-organ failure. Although some secondary endpoints could not be collected due to COVID-19 restrictions, recruitment and retention rates were high. Variability in glycaemic control improved, and differences between arms in terms of hypogylycaemia, insulin dosing and glucose variability all point towards a favourable metabolic profile and ketone bodies being used as a substrate preferentially over glucose. In keeping with tissue ketone body metabolism, ketosis was not associated with development of arterial blood acidaemia. Exploratory untargeted metabolomic analyses showed clear separation of arms at the end of the intervention. Metabolite abundance data suggest that a favourable metabolic profile developed in response to the intervention, a hypothesis that requires prospective testing in a larger trial.

Prior to this trial, concerns had been raised regarding safety and feasibility of inducing ketosis in physiologically unstable patients. Ketosis is traditionally associated with pathological states in clinical medicine^47^. However, staff and patient education and engagement led to excellent rates of recruitment and retention of patients. Staff questionnaires suggest not only a high level of enthusiasm for the study, but also that this was, in the main, scalable, if a pre-made enteral feed could be sourced, of which several are available commercially.

Support for the study may be in part due to the safety profile observed, with little differences seen in adverse events or tolerability, except for incidence of diarrhoea. We had not pre-specified the definition of diarrhoea, and several such definitions exist. Diarrhoea is common in critically ill patients^22^, and it is not clear if the high medium chain triglyceride load led to an increased incidence although diarrhoea is a well-known consequence of enteral feeding when there is a high proportion of medium chain triglycerides^48^. Future trials should pre-specify such definitions for monitoring purposes.

Octanoic acid and decanoic Acid were delivered as part of the intervention, and the increased presence of these in the circulation supports data from recent stable isotope studies that gastrointestinal absorption in not a limiting factor^49^. Moderate levels of both medium chain fatty acids and ketone bodies in the participants receiving ketogenic enteral feeding suggest that the pathway of medium chain triglyceride lipolysis, absorption, hepatic ketogenesis and extra-hepatic ketone body utilisation is functionally intact and operative in these patients, supporting the hypothesis that provision of a ketogenic enteral feed to critically ill patients provides an alternative substrate. Dodecanoic acid is not present in either the ketogenic or standard enteral feeds and was no different between arms, acting as a form of internal control. Despite the altered composition of the novel enteral feed formula, nutritional adequacy was achieved equally across arms. In an exploratory analysis the respiratory quotient (RQ) was lower in the ketogenic enteral feeding arm, adding further data suggestive of ketone metabolism occurring for energy generation^50^. While the significance of the equivalence in pyruvate concentrations is unclear, the lower lactate levels seen in the intervention arm are also suggestive of substrate switching.

The collection of outcome data was significantly impacted by the COVID-19 pandemic. Bedside collection of 24-hour urinary data (and hence nitrogen levels) and reliable muscle ultrasound measurements were not possible due to a combination of infection control requirements, the use of PPE, and staff shortages due to redeployment. Routinely collected physical outcome data were limited to the minimum possible as a result. Staff acceptability of the study protocol indicated some level of difficulty with collecting the quality-of-life data both retrospectively (pre-ICU admission) and following ICU discharge. This is highly likely to be related to difficulties in communicating with family members due to COVID-19-related suspension of hospital visiting.

Of note, measurement of muscle mass was neither an essential nor recommended outcome measure in the Core Outcome Set for metabolic and nutritional interventions in critically ill patients and would therefore be unlikely to form part of our subsequent efficacy trial^51^.

Exploratory metabophenotyping demonstrated good metabolic separation between arms following the 10-day intervention. Given the multiple tissue metabolites contributing to plasma metabolite profiles, this separation lends weight to the hypothesis that ketone bodies are being used for substrate metabolism in diverse physiological pathways. Pathways unrelated to metabolism of nutritional lipids were differentially regulated implying tissue metabolism was additionally altered. While suggestive of an advantageous metabolic profile, these data should be viewed as somewhat preliminary, given the small patient numbers. Alterations in Cahill cycle flux result in a differential metabolite abundance in the alanine pathway, suggesting a decrease in muscle protein breakdown for alanine production^10^. Pentose and glucoronate interconversions suggest a differential regulation of the bioenergetic state^52^, but no conclusions can be drawn regarding improvement of bioenergetic failure from this small sample.

### Strengths and Limitations

This study has considerable strengths. First, the very comprehensive prospective data collection gives a high level of confidence in the safety data for future trials, and suggests that such trials are unlikely to require such extensive data collection (thus reducing the data collection burden). Second, the extensive biochemical and metabolomic analysis gives insight into metabolic processes brought into play by the delivery of ketogenic enteral feeding. Limitations include the need to focus on safety and feasibility which determined the sample size. Imbalances in the study cohort between arms are likely driven by this (e.g., APACHE score inequality versus SOFA equality). Regardless, all patients recruited were in multi-organ failure, and at risk of both altered substrate utilisation, muscle wasting, and subsequent physical functional impairment. All signals reported relating to efficacy should be viewed as hypothesis generating only as this trial was not powered to detect this. A further limitation is the missing data for some of the physical outcomes – an indication of the strain put on healthcare systems by the COVID-19 pandemic. From a feasibility perspective, future trials would require dedicated time and funding for this, as these data expose the fragility of using routinely collected physical outcome data.

## Conclusions

Ketogenic feeding in critically ill patients with multi-organ failure is safe and feasible. Patients who received ketogenic enteral feeding developed a different metabolic profile from controls. The efficacy of this altered metabolic profile in improving patient outcomes requires testing in prospective trials.

## ACKNOWLEDGEMENTS

AM, DB, SE, KR, HM and ZP received a Research for Patient Benefit grant (PB-2006) from the National Institute for Health Research (NIHR). HM received funding from the NIHR’s Biomedical Research Centre (BRC) at University College London Hospitals, London, UK. The research was also supported by the NIHR BRC based at Guy’s and St Thomas’ NHS Foundation Trust and King’s College London and by the NIHR BRC at Great Ormond Street Hospital. The views expressed are those of the authors and not necessarily those of the NHS, the NIHR, the Department of Health or the funders. Metabolomic data was supported by an educational grant from Nestle Health Sciences. Vitaflo International Ltd were involved in initial discussions about the study and provided the K.Quik® component for the ketogenic feed gratis. Neither Vitaflo International Ltd nor Nestle Health Sciences contributed to study design, study implementation, data analysis or interpretation.

We would like to thank the patients (and their families) who took part, and the research nurses of both recruiting centres for their willingness to engage. Specifically: Maria Fernandez, Filipa Santos, Amaia Garcia, Fatima Seidu, Katie Sweet. We would also like to thank those who funded this study (below).

## Author contributions

AM, DB,SE,KR,HM and ZP conceived and designed the clinical trial.

AM, DB,AL, KR, ZP, JP, RP, AP, TM, FS, FS1, KL carried out and delivered the clinical trial.

AM, ZP, PA, DW, HC, IA, SE, TB, SH performed the analyses.

All authors read and approved the manuscript.

