## Supplementary Information for "Alternative Substrates in the Critically Ill Subject (ASICS): Safety, Feasibility, Tolerability and Metabolic Profiling of a Novel Ketogenic Feed"

Angela McNelly[1], Anne Langan[2], Danielle E. Bear[3,4,5], Alexandria Page[6], Tim Martin[6], Fatima Seidu[6], Filipa Santos[6], Kieron Rooney[7], Kaifeng Liang[1], Simon J Heales[9], Tomas Baldwin[8], Isabelle Alldritt[10], Hannah Crossland[10], Philip J. Atherton[10], Daniel Wilkinson[10], Hugh Montgomery[11,12], John Prowle[1,6], Rupert Pearse[1,6], Simon Eaton[8] and Zudin A. Puthucheary[1,6,*]

*[1]William Harvey Research Institute, Faculty of Medicine & Dentistry, Queen Mary University of London, [2]Department of Dietetics, Adult Critical Care Unit, Royal London Hospital, London, [3]Department of Nutrition and Dietetics St Thomas' NHS Foundation Trust, [4]Department of Critical Care, Guy's and St. Thomas' NHS Foundation & King's College London (KCL) NIHR BRC, London, [5]Kings College London, [6]Adult Critical Care Unit, Royal London Hospital, London, [7]Department of Critical Care, Bristol Royal Infirmary, Bristol, [8]Developmental Biology & Cancer, UCL Great Ormond Street Institute of Child Health, London, [9]Genetic & Genomic Medicine Department, UCL Great Ormond Street Institute of Child Health, London, [10]Centre of Metabolism, Aging & Physiology (COMAP), MRC-Versus Arthritis Centre for Musculoskeletal Aging Research & NIHR Nottingham BRC, University of Nottingham, Nottingham, [11]University College London (UCL), [12]UCL Hospitals NHS Foundation Trust (UCLH), National Institute for Health Research (NIHR) Biomedical Research Centre (BRC), London.*

***CORRESPONDENCE TO**
Dr Zudin Puthucheary
Critical Care and Perioperative Medicine Research Group,
Adult Critical Care Unit,
Royal London Hospital,
London, E1 1BB
United Kingdom


**METHODS:**

### 1. Feeding regimens

#### *1.1 Nutritional procedures and individualised energy and protein targets*

Nasogastric (NG) tubes were inserted as part of routine clinical care for mechanically ventilated patients. Following confirmation of correct tip position, according to national standards[1], continuous enteral feeding with either standard feed (according to local Trust protocols) or ketogenic feed was commenced.

Once reviewed by the ICU dietitian, feed volumes and compositions were adjusted according to individualised targets for energy and protein. Energy targets were mostly determined using the modified Penn State Equation (using actual body weight), the most accurate when compared to indirect calorimetry in critically ill adults[2]. Weight-based equations (e.g., 25 kcal/kg) could be used where considered more appropriate, but indirect calorimetry was not available routinely. A minimum protein target of 0.83 g/kg/day was used with adjustments for other clinical reasons (e.g., use of continuous renal replacement therapy). Ideal or adjusted body weights were used where appropriate (e.g., high body mass index) as decided by the dietitian.

#### *1.2 Feed management for procedures*

Where required for clinical reasons (e.g., airway management), feed was stopped according to local guidelines and restarted as soon as possible following the procedure. Where a ‘feed free’ time was required for enteral drug administration (e.g., phenytoin), an appropriate feeding regimen was determined by the dietitian.

#### *1.3 Management of high gastric residual volumes*

A Gastric Residual Volume (GRV) threshold of 300mls was set. Following administration of metoclopramide and/or erythromycin, and alleviation of any other factors which might have been reducing gastric absorption or causing ileus (e.g., drug use which could be discontinued), high GRV was managed according to local guidelines.

### 2. Recruitment

Potential participants were screened by research nurses and recruited by a member of the research team.

#### *2.1 Exclusion criteria*

These included primary neuromyopathy or significant neurological impairment at the time of ICU admission that would preclude physical activity; unilateral/bilateral lower limb amputation; requirement for sole or supplementary parenteral nutrition; need for specialist nutritional intervention; known inborn error of metabolism; participation in another clinical trial. Patients at risk of refeeding syndrome (based on NICE guidelines) were assessed on an individual basis[1].

### 3. Randomisation and blinding

#### *3.1 Procedures*

Randomisation by a member of the research team took place once assent was obtained; feeding commenced as soon as possible after randomisation (according to standard clinical timing). An independent remote electronic web-based random allocation service was used to generate an unpredictable treatment group allocation and to conceal that outcome from the members of the research team until assignment occurred. Investigators performing muscle ultrasound undertook inter-observer variability assessments after training and prior to the trial commencing. All images taken during the trial were allocated an anonymised code. The off-site investigator (DB) analysing the muscle ultrasound scans was blinded to the feeding regimen allocation until post-analysis. Likewise, the majority of secondary outcomes (e.g., length of stays and days of ventilation) were collected from paper or electronic systems not susceptible to bias.

#### *3.2 Allocation imbalance management*

To correct an imbalance in numbers per arm, ethical approval was obtained so that randomisation could continue until 37 patients in total had been recruited.

### 4**. Criteria for Premature Withdrawal**

In the event of the attending consultant or critical care dietitian having any clinical concerns relating to a patient in the ketogenic arm, a switch to standard feeding occurred. Feeding guidelines included the use of prokinetic drugs, but ultimately their use was at the discretion of the treating clinicians, as is the case for routine clinical care. In the event that a protocol deviation occurred, data continued to be accrued to inform future studies. Protocol violations included meeting less than 80% of prescribed energy and protein over the period of the study protocol (10 days), cross-over between study arms, and need for parenteral nutrition or post-pyloric feeding.

**5. Secondary Endpoints**

Additional secondary endpoints included functional outcomes (number of days to first sit-to-stand test and to first bed-to-chair transfer prior to ICU discharge; 6-Minute Walk Test and Short Physical Performance Battery at ICU/hospital discharge); clinical data (blood biochemistry; length of stay on ICU and in hospital; discharge location; number of days of mechanical ventilation; infection); and metabolomic analysis. Follow-up at 3-, 6-, and 12-months included Health-Related Quality of Life (EQ-5D-5L questionnaire), employment status and primary health care usage costs (Table 1, main paper).

*5.1 Blood and Urine sample preparation and analysis*

Blood samples (in EDTA tubes) and urine samples (in universal tubes) were centrifuged at 2500 g for 5 min and the protein-free supernatant removed and stored at -80°C, before analysis for plasma medium chain fatty acids, lactate and beta-hydroxybutyrate, and urinary beta-hydroxybutyrate, respectively, by gas chromatography/mass spectrometry[3].

For acetoacetate and pyruvate, which are not stable in plasma, blood samples were taken on trial days 1, 7 and 10. At the recruiting site, 1.5-2 ml of whole blood was added to a pre-weighed plastic tube containing 5 ml of 0.77 mol/l perchloric acid previously cooled to 0° C. After mixing, the tubes were re-weighed, centrifuged at 2500 g for 5 min and the protein-free supernatant removed and stored. The dilution of the blood was determined by the weight changes measured. Samples were stored at -80°C until analysis. 100ul supernatant plus 20ul internal standard mix (100uM $^{13}C_3$-pyruvate, $^{13}C_4$-acetoacetate) were added to 150ul *O*-(2,3,4,5,6-Pentafluorobenzyl)hydroxylamine hydrochloride (PFHBA) in 1M HCl. After 1 hour at room temperature, 50ul concentrated $H_2SO_4$ was added to remove PFHBA, then 1ml $H_2O$, plus 3ml ethyl acetate. The organic phase was transferred to a new tube, and 1ml 0.2N $H_2SO_4$ added to remove any residual PFHBA. The organic phase was again removed, evaporated under $N_2$. 100ul *N*-Methyl-*N*-trimethylsilyltrifluoroacetamide/1% Chlorotrimethylsilane was added, plus 50ul pyridine. After incubation at 75°C for 1 hour, samples were analysed by GC/MS (equipment as above), inlet temperature 250°C, helium flow rate 1.5mL/min, 2µl injection and 1:10 split ratio. Oven temperature gradient was 100°C, held for 1minute and then ramped to 190°C at 5°C/min, then to 300°C at 40°C/min. Compounds were analysed by negative chemical ionization (methane flow 2mL/min). The following fragment ions were detected in selected ion monitoring mode: m/z 174 (pyruvate), 177 ($^{13}C_3$-pyruvate), 98 (acetoacetate), 102 ($^{13}C_4$-acetoacetate. Concentrations were corrected for blood dilution in perchloric acid.

*5.2 Ultrasound image acquisition training*

*5.2.1 Trial personnel training*

All trial personnel involved in image acquisition were trained onsite by a researcher experienced in the method (AM or ZAP). Personnel then underwent a period of practice and were expected to confirm inter- and intra-rater reliability in 10 healthy subjects at their local sites. Sites were deemed trained and ready to begin recruitment if images analysed independently (ZAP) were found to have an Intraclass Correlation Coefficient >0.9. However, during the pandemic training was not performed in protective equipment, and new staff did not have the same level of training since access to healthy controls was limited, as was time to train and determine the Intraclass Correlation Coefficient.

*5.2.2 Machines used*

Measurements were made using the following ultrasound machines: Sonosite M-Turbo, C60XI 5-20MHz probe (FijiFilm SonoSite Ltd, London, UK) [Royal London Hospital]; Sonosite M-Turbo machine HFL 50x/15-6MHz transducer [Bristol Royal Infirmary].

*5.2.3 Image acquisition*

The method for measurement was as that described previously[4-6]. Three images were captured at each time point.

*5.2.4 Image analysis*

Images were stored on encrypted memory sticks under a pseudo-anonymized patient number and transferred to password-protected computers. Images were then analysed offline by a single experienced member of the research team (DB) not involved with the clinical management of any study patients and blinded to the intervention they received (Image J software, National Institutes of Health, US). Rectus femoris muscle cross sectional area was taken as the average of three consecutive measurements within 10% of one another. Scans from 1 in 4 patients underwent re-analysis to ensure good intra-rater agreement.

**6.2 Metabolomic Analysis**

Blood samples (in lithium heparin tubes) were centrifuged at 2500 g for 5 min and the protein-free supernatant removed and stored at -80°C before transfer for metabolomic analysis.

*6.2.1 Liquid chromatography*

Separation was performed on an Accela UHPLC pump and autosampler. For polar metabolites, an InfinityLab Poroshell 120 HILIC-Z column (2.1mm x 150mm x 2.7µm, Agilent) was used with mobile phase A 10mM ammonium formate in 90% acetonitrile with 0.1% formic acid and mobile phase B 10mM ammonium formate in 50% acetonitrile with 0.1% formic acid. For non-polar metabolites, a

Zorbax SB-Aq RRHD column (2.1mm x 100mm x 1.8µm, Agilent) was used with mobile phase A dd$H_2O$ with 0.1% formic acid and mobile phase B methanol with 0.1% formic acid.

| Time | Mobile Phase A (%) | Mobile Phase B (%) |
|---|---|---|
| **HILIC (flow rate 400µl/min)** | | |
| **0.00** | 99.00 | 1.00 |
| **1.00** | 99.00 | 1.00 |
| **3.00** | 85.00 | 15.00 |
| **6.00** | 5.00 | 95.00 |
| **10.00** | 5.00 | 95.00 |
| **10.50** | 99.00 | 1.00 |
| **15.50** | 99.00 | 1.00 |
| **Reverse phase (flow rate 400µl/min)** | | |
| **0.00** | 99.00 | 1.00 |
| **0.50** | 99.00 | 1.00 |
| **2.00** | 50.00 | 50.00 |
| **10.00** | 1.00 | 99.00 |
| **12.00** | 1.00 | 99.00 |
| **12.50** | 99.00 | 1.00 |
| **17.50** | 99.00 | 1.00 |

**Table S1: Gradient conditions and flow rates for liquid chromotography.** HILIC= Hydrophilic interaction chromatography

*6.2.2 Mass spectrometry*

Data acquisition was performed by a Q-Exactive high resolution mass spectrometer (Thermo Fisher Scientific) operated in the positive and negative ionisation modes.

| Parameter | HILIC | | Reverse phase | |
|---|---|---|---|---|
| **Mode** | Positive | Negative | Positive | Negative |
| **MS scan parameters** | | | | |
| **Scan type** | Full MS | | Full MS | |
| **Scan range** | 70 – 1,050 m/z | | 150 – 2,000 m/z | |
| **Fragmentation** | None | | None | |
| **Resolution** | 70,000 | | 70,000 | |
| **AGC target** | 1e6 | | 3e6 | |
| **Maximum IT** | 100ms | 200ms | 100ms | 250ms |
| **Microscans** | 5 | | 1 | |
| **Sheath gas flow** | 54 | | 48 | |
| **Aux gas flow** | 13 | | 11 | |
| **Sweep gas flow** | 0 | 3 | 5 | |
| **Spray voltage** | 4kV | 3.5kV | 3kV | 3.5kV |
| **Capillary temperature** | 280°C | 320°C | 300°C | 320°C |
| **S-lens RF level** | 50 | 60 | 60 | |
| **Aux gas flow heater temperature** | 430°C | 320°C | 300°C | |
| **MS/MS scan parameters** | | | | |
| **Resolution** | 17,500 | | 17,500 | |
| **AGC target** | 1e6 | | 1e6 | |
| **Maximum IT** | 50ms | | 50ms | |
| **topN peaks** | 3 | | 3 | |
| **Isolation window** | 1.5 m/z | | 1.5 m/z | |
| **Normalised collision energy** | 25, 60, 100% | | 25, 60, 100% | |

**Table S2: Mass Spectrometry parameters for positive and negative ionisation modes.** HILIC= Hydrophilic interaction chromatography

*6.2.3 Sample pre-processing*

Pre-processing was performed in R using XCMS package[7]. Scripts are available on request. Chromatograms were extracted using the centWave algorithm at a tolerance of 20 parts per million (ppm). Chromatograms were aligned using the Obiwarp method with bin size 0.6. Bandwidth was determined manually for correspondence. Missing values were imputed via integration of peak area. For each ion mode and polarity, data were exported as a data frame of metabolite feature *vs* sample ID with associated chromatographic peak areas for each detected metabolite.

Following recommended guidelines, metabolite features were retained when: peaks were present in at least 70% of pooled QC samples, relative standard deviation was less than 30%, and the extraction blank to mean QC peak area was less than 50%[8]. Probabilistic quotient normalisation was applied to the remaining features, then missing values were imputed using the k-Nearest Numbers algorithm. Data were log transformed prior to analysis.

**7. Safety and Adverse Event Management**

Where an adverse event was considered serious (SAE) or unexpected (i.e., not listed in the protocol as an expected occurrence), the Principal Investigators at both sites were mandated to report it to the Chief Investigator, who had responsibility for informing the Sponsor's research and development department within 24 hours and the main Research Ethics Committee (REC) within 15 days using the REC's standard template. All other Adverse events (AEs) were reported to Barts Health NHS Trust.

*7.1 Definitions*

• Adverse Events are any untoward medical occurrence or effect in a patient participating in the trial, which does not necessarily have a causal relationship with trial treatment. An Adverse Event can therefore be any unfavourable symptom or disease temporally associated with the use of the trial treatment, whether or not it is related to the allocated trial treatment.

• Serious Adverse Events (SAEs) are AEs that

  o Result in death;

  o Are life-threatening;

  o Require in-patient hospitalisation or prolongation of existing hospitalisation;

  o Result in persistent or significant disability/incapacity; or

  o Result in a congenital anomaly/birth defect

• All AEs were graded for severity:

0. None: indicates no event or complication.

1. Mild: complication results in only temporary harm and does not require clinical treatment.

2. Moderate: complication requires clinical treatment but does not result in significant prolongation of hospital stay. Does not usually result in permanent harm and where this does occur the harm does not cause functional limitation to the patient.

3. Severe: complication requires clinical treatment and results in significant prolongation of hospital stay, permanent functional limitation.

4. Life-threatening: complication that may lead to death.

5. Fatal: indicates that the patient died as a direct result of the complication/adverse event.

*7.2 Potential "expected" AE's in patients with multi-organ failure*

- Abdominal distension – new, clinically significant change in appearance. Considered severe if acute obstruction.
- Abdominal pain – new, localised to abdomen and requiring more than just simple analgesia. Considered severe if not controlled with opiates.
- Electrolyte disturbance – new change that is clinically significant requiring active monitoring or treatment.
- Hypersensitivity reaction (anaphylactic reaction) – anaphylactic reaction.
- Hypoglycaemia – new, clinically significant hypoglycaemia requiring active monitoring or treatment.
- Ischaemic bowel – inferred on radiology or diagnosed visually, e.g. surgery or endoscopy.
- Nausea requiring treatment with anti-emetics; Vomiting – any episode
- Regurgitation/aspiration – any episodes

**8. Data Collection**

Routinely measured physiological data were collated through the 10-day study period. Medications administered to the patient each day were recorded, as were details of energy and protein prescribed and received (from feed, propofol and additional glucose), enabling total calories and grams of protein delivered, the percentage of target calories and protein delivered, and nutritional delivery to be compared between the two arms. Where possible functional milestones (days before Bed-to-Chair transfer and Sit-to-Stand times; 6-Minute Walk Distance and Short Physical Performance Battery) were collected at ICU/hospital discharge. Length of mechanical ventilation, ICU and hospital stays, and hospital discharge destination were collected and noted from medical records. Follow-up data (health-related quality of life; employment and primary care usage) were collected at 3-,6- and 12-months post-ICU discharge if relevant staff were available given COVID-19-related limitations. Data were collected initially into a paper Case Report Form and then transferred in anonymised form to a secure database.

**Results**

*Recruitment and Retention*

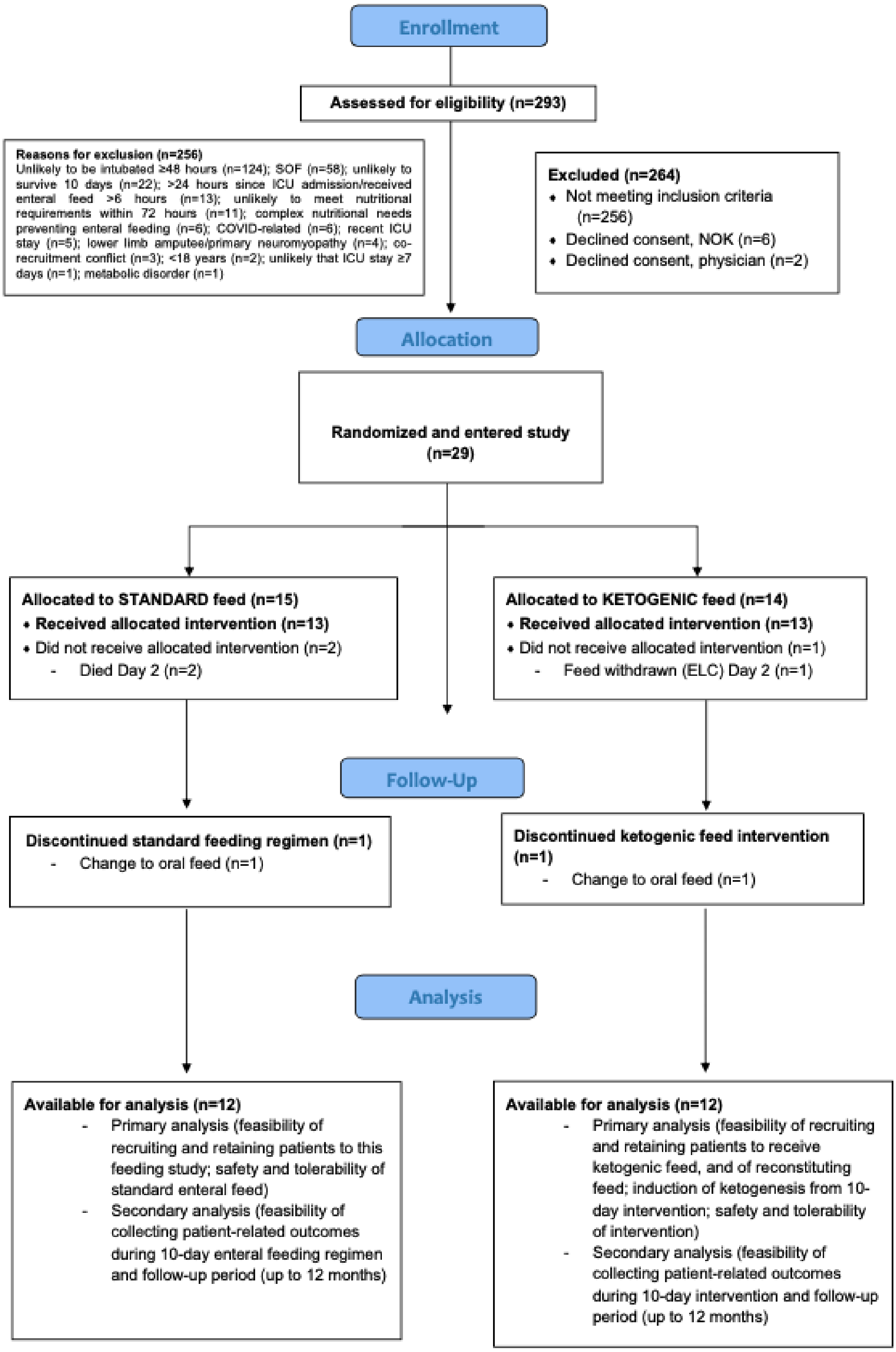


**Figure S1: Consort flow diagram**

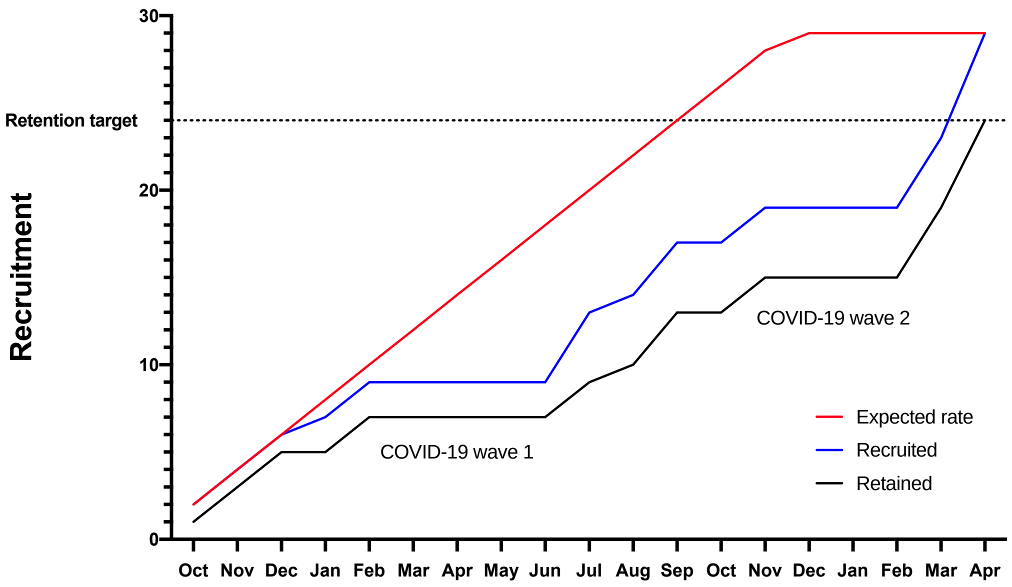


**Figure S2: Recruitment rates and the effects of COVID-19 waves during the pandemic**

| Site | No. Screened | No. Recruited (%) | No. Retained (%) |
|---|---|---|---|
| **RLH** | 258 | 27 | 23 |
| **BRI** | 35 | 2 | 1 |
| **TOTAL** | 293 | 29 (9.9) | 24 (8.2) |

**Table S3: Recruitment across both sites.** No.=number; RLH=Royal London Hospital BRI=Bristol Royal Infirmary

| Withdrawal reason | Ketogenic Feed Arm (n=14) No. (%) | Standard Feed Arm (n=15) No. (%) |
|---|---|---|
| **ICU d/c<7d (died)** | 0 | 2 |
| **Feed withdrawn (ELC)** | 1 | 0 |
| **Feed change to oral** | 1 | 1 |
| **Total** | 2 (14.3) | 3 (20) |

**Table S4: Reasons given for premature withdrawal of participants.** ICU= Intensive Care Unit; d/c<7d=discharged in less than 7 days; ELC=End of life care.

*Acceptability and Feasibility*

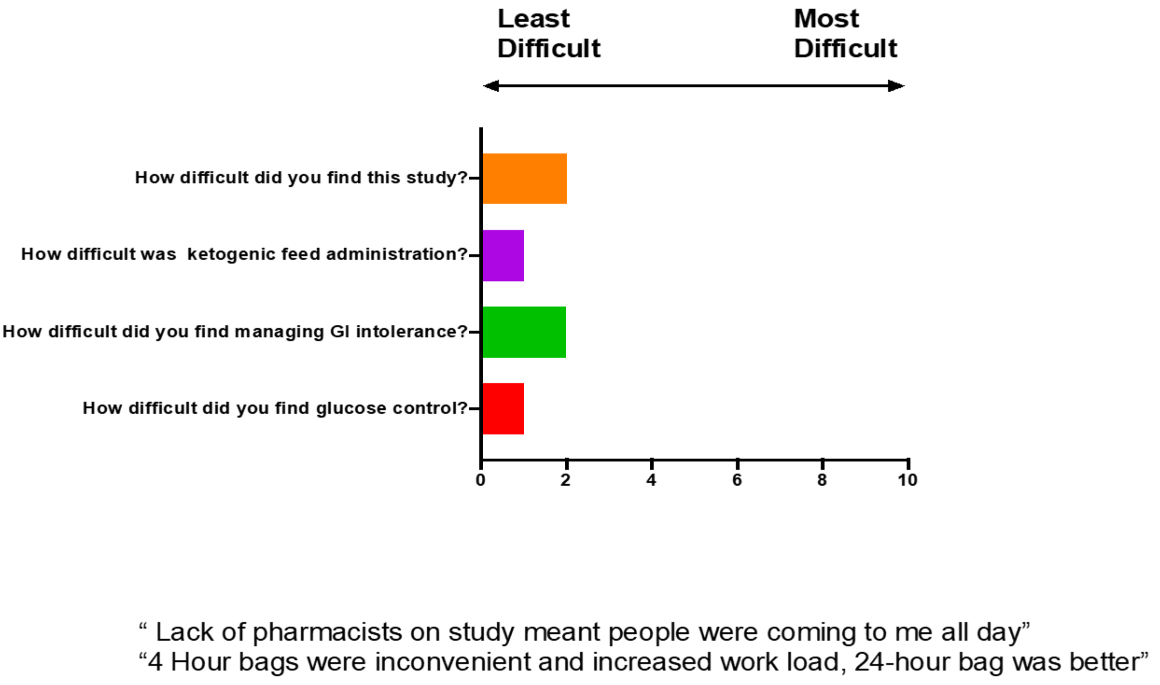


**Figure S3: Staff questionnaire data on feasibility and acceptability of delivering the intervention.**

*Safety data*

| Serious Adverse Event summary | Outcome |
|---|---|
| **Brain stem death due to trauma-related dissection of carotid arteries** | Unrelated |
| **Death from pulmonary embolism and biventricular failure** | Unrelated |
| **Metabolic acidemia secondary to hyperchloremia and dapaglifozine** | Unrelated; Clinicians stopped dapaglifozine and continued ketogenic feed |
| **Death from chest sepsis** | Unrelated |

**Table S5: Severe Adverse Events and trial related outcomes.**

| **Days/events reported, No. (% total days/events)** | **High GRV*** | **Vomiting**** | **Diarrhoea (3-days)[a]** | **Diarrhoea (daily)[b]** | **Prokinetic Use** | **Hypoglycaemia^** | **Hyper-glycaemia^^** |
|---|---|---|---|---|---|---|---|
| **STANDARD ENTERAL FEED (n=15)** | 25(19.8) | 7 (5.5) | 8 (53.3) | 54 (42.9) | 29 (22.8) | 2 (1.6) | 73 (57.5) |
| **KETOGENIC ENTERAL FEED (n=14)** | 19 (17.3) | 9 (8.1) | 10 (76.9) | 54 (53.5) | 23 (20.7) | 0 (0.0) | 29 (26.9) |

**Table S6**: **Adverse events** *Days with Gastric Residual Volume ≥300mls; ^Blood glucose ≤3.9mM; **Days with any vomiting (>10mls); ^^Blood glucose ≥10mM; [a]Three-day episodes of Bristol Stool Score T6 or T7; [b]Days with Bristol Stool Score T6 or T7. GRV=Gastric Residual Volume.

| | **Events reported, No. (%)** | | **Mean (95% CI)** | |
|---|---|---|---|---|
| | AKI | Metabolic acidosis | Base Excess | Bicarbonate |
| **STANDARD ENTERAL FEED** | 1 (6.6) | 0 (0.0) | 1.78 (2.61-0.95) | 26 (16.8-25.2) |
| **KETOGENIC ENTERAL FEED** | 1 (7.1) | 2 (14.3)[#] | -1.68 (-1.01- -2.35) | 22.6 (23.3-21.9) |

**Table S7: Adverse events** #Deemed by clinical team to be related to the ketogenic feed. AKI=Acute Kidney Injury.

*Urinary Ketone Body Concentration*

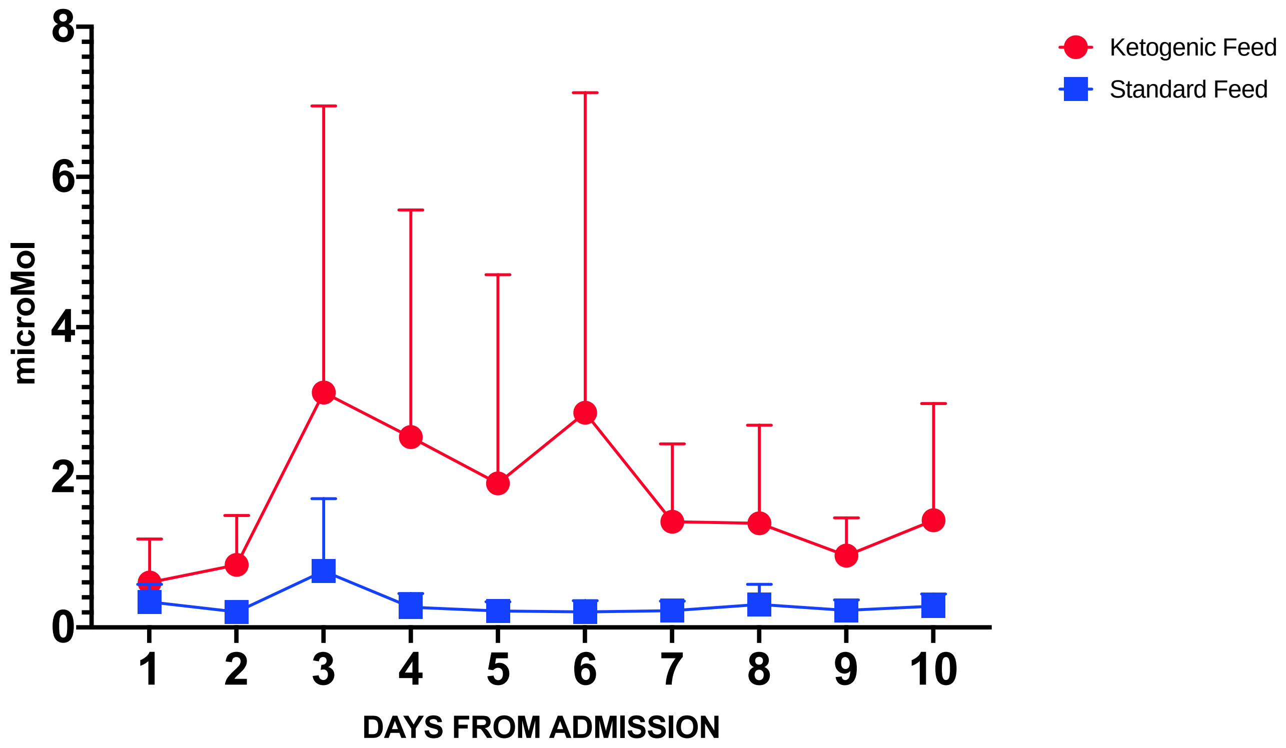


**Figure S4: Urinary beta-hydroxybutryrate during the 10-day intervention. Red lines represent ketogenic feeding, and blue lines controls.**

*Plasma Fatty Acid Concentrations*

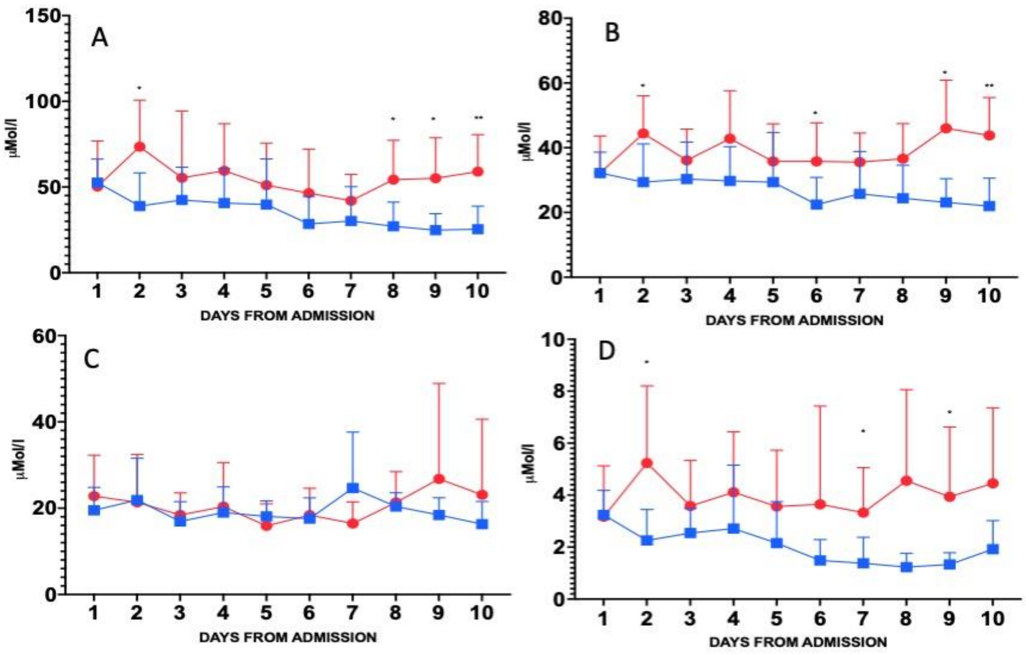


**Figure S5: Octanoic acid (A), decanoic acid (B), and dodecanoic acid (C) in plasma during the 10-day intervention; Octanoic-to-Dodecanoic Acid ratio (D).** *p<0.05, **p<0.01 between arms (Mann Whitney-U test).

*Respiratory Quotients*

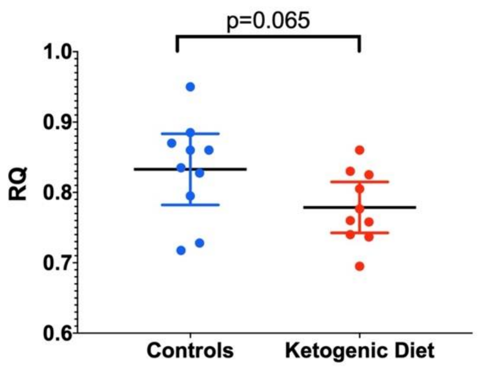


**Figure S6: Respiratory Quotient (RQ) of a sub-group of patients receiving the ketogenic (red) and control (blue) feeds.**

*Metabophenotyping*

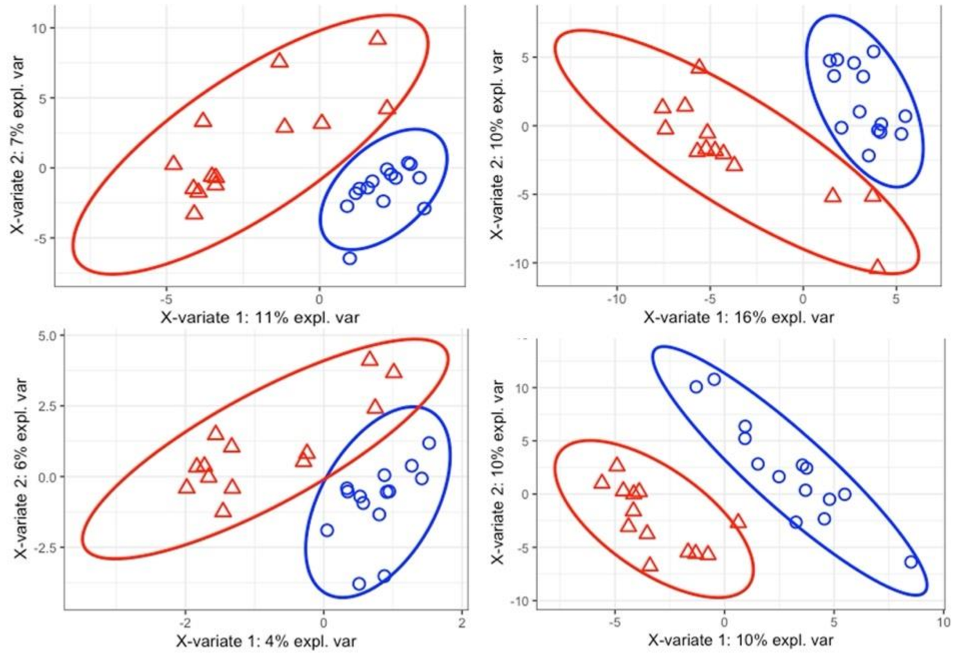


**Figure S7: Sparse Partial Least Squares Discriminant analysis of patient randomised to ketogenic feeding on Day 1 (red triangle) and control feeding on day 1 (blue sphere). Clockwise from top right: polar positive, non-polar positive, non-polar negative, polar negative.** Error rates are >20% (17%, 41%, 31% and 20% respectively), suggesting plot is overfitted and not true variance.

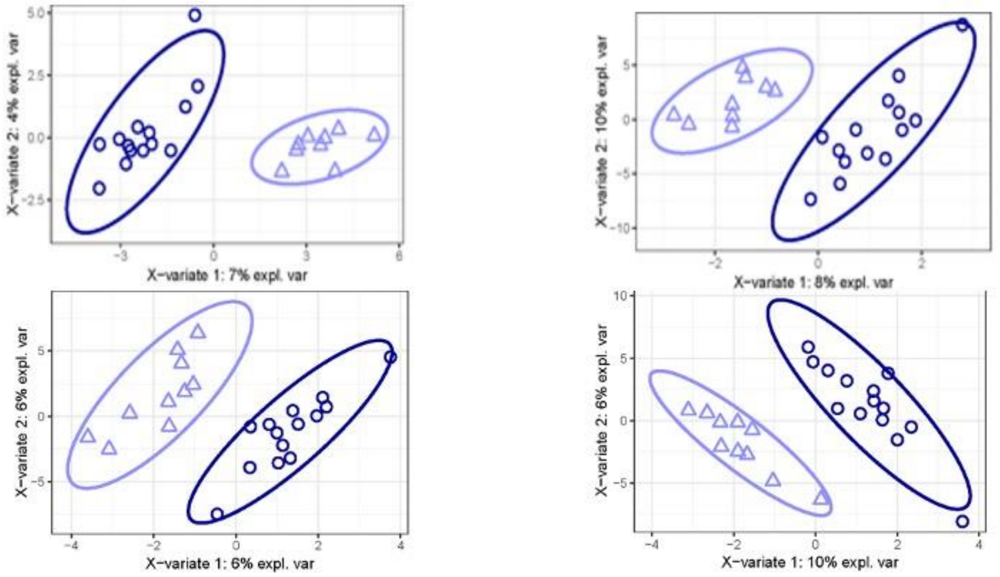


**Figure S8: Sparse Partial Least Squares Discriminant analysis of patient randomised control feeding on Day 1 (blue sphere) and day 10 (blue triangle). Clockwise from top right: polar positive, non-polar positive, non-polar negative, polar negative.** Error rates are mixed >20% (17%, 25%, 15% and 25% respectively), suggesting plot is at risk of overfitting a opposed to true variance.

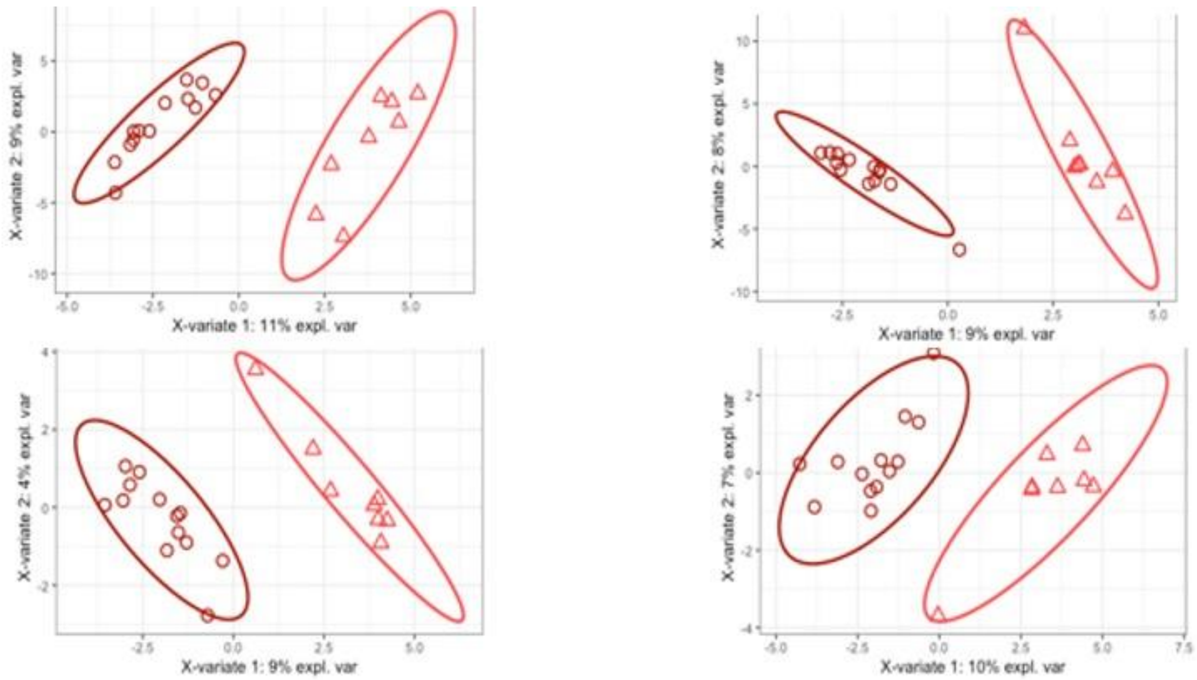


**Figure S9: Sparse Partial Least Squares Discriminant analysis of patient randomised to ketogenic feeding on Day 1 (red sphere) and day 10 (red triangle). Clockwise from top right: polar positive, non-polar positive, non-polar negative, polar negative.** Error rates are <20% (10%, 12%, 15% and 10% respectively), suggesting plot is result of true variance.
